# Multimodal wearable sensors inform cycles of seizure risk

**DOI:** 10.1101/2022.07.10.22277412

**Authors:** Nicholas M. Gregg, Tal Pal Attia, Mona Nasseri, Boney Joseph, Philippa J. Karoly, Jie Cui, Rachel E. Stirling, Pedro F. Viana, Thomas J. Richner, Ewan S. Nurse, Andreas Schulze-Bonhage, Mark J. Cook, Gregory A. Worrell, Mark P. Richardson, Dean R. Freestone, Benjamin H. Brinkmann

**Author notes:** Corresponding author: Nicholas Gregg, Bioelectronics Neurophysiology and Engineering Laboratory, Department of Neurology, Mayo Clinic, Rochester, MN 55905, USA.

## Abstract

**Objective:** Seizure unpredictability is a major source of disability for people with epilepsy. Recent work using chronic brain recordings has established that for many individuals with epilepsy seizure risk is not random, but corresponds to circadian and multiday (multidien) cycles in brain excitability. Here, we aimed to evaluate whether multimodal wearable device recordings can characterize cycles of seizure risk, and compare wearables performance with concurrent chronic brain recordings.

**Methods:** Fourteen subjects underwent long-term ambulatory monitoring with a multimodal wrist worn device (measuring heart rate, heart rate variability, accelerometry, tonic and phasic electrodermal activity, temperature) and an implanted responsive neurostimulation system (measuring interictal epileptiform abnormalities (IEA) and electrographic seizures). Wavelet time-frequency analyses identified circadian and multiday cycles in wearable and brain recordings. Circular statistics assessed seizure phase locking to cycles in physiology.

**Results:** Ten subjects met inclusion criteria. The mean recording duration was 232 days. Seven subjects had reliable electrographic seizure detections (mean 76 seizures). Seizure phase locking to multiday cycles occurred in six (IEA), five (temperature), four (heart rate, phasic electrodermal activity), and three (accelerometry, heart rate variability, tonic electrodermal activity) subjects. Seizure phase locking to residual HR multiday cycles (HR after regression of correlated physical activity (ACC)) increased to six subjects.

**Interpretation:** Long timescale cyclical changes in wearable recordings are common in epilepsy, and seizures occur at preferred phases of these cycles for many individuals. Broadly accessible wearable technology can provide new insights into the chronobiology of epilepsy with implications for seizure forecasting.

## Introduction

Work using chronic brain recordings from humans,^1-4^ dogs,^5,6^ and mice^7^ has recently established that seizure risk fluctuates over daily (circadian) and multiday (multidien) cycles for many individuals with epilepsy. Pioneering work from the early 20^th^ century identified circadian and multiday^8-10^ periodicities in seizure occurrences based on meticulous clinical records and seizure diaries kept for individuals living in supervised care facilities. More recently, chronic intracranial recordings from clinical^4,11-14^ and investigational^1,2,5,6,15^ devices have revealed that for many individuals with epilepsy interictal epileptiform activity (IEA) is modulated over circadian and multiday cycles, and that seizures occur at preferred phases of these underlying cycles.^3^ By measuring seizure risk relative to physiological signals amenable to continuous temporal sampling (i.e. IEA), one may achieve superior characterization of seizure risk compared to risk defined by fixed clock-time based cycle periods,^16^ with implications for seizure risk forecasting.^13^ These long timescale fluctuations in IEA and seizure risk may reflect long-term changes in brain excitability.^17-20^

This body of work has provided important insights into epilepsy physiology, with relevance for clinical care. However, the requirement of chronic brain recordings is a barrier to widespread accessibility. Recent work using data from a wearable fitness device^21^ provides exciting evidence that long timescale changes in physiology (heart rate (HR)) may also inform seizure risk cycles. The mechanisms behind the association of seizure risk and multiscale HR cycles in epilepsy are unclear, but may include interactions between brain excitability and autonomic regulation,^21-23^ in addition to potential behavioral and homeostatic mechanisms.

Circadian and multiday cycles in human physiology have been described in normal health and disease^24-31^. Of particular interest multiday cycles, including circaseptan^32^ (weekly) and circamonthly^33^ cycles, have been identified in the regulation of immune,^26^ endocrine,^28,34^ metabolic,^35^ and cardiovascular^36,37^ systems, in human behavior,^32^ as well as in brain excitability^17,20^ and seizure risk.^2-6,12^ Prior work on naturally occurring human and canine epilepsy has established that multiday cycles are common, with evidence for group-level circa-weekly, bi/tri-weekly, and monthly cycles.^12^ Wearable devices that provide multimodal recordings of body physiology are available to the lay population in popular commercial smartwatch systems, as digital health has become a growth area for the technology sector. This trend is likely to continue as wearable sensors become more popular, additional sensor modalities are integrated into commercial systems, and near real-time cloud-based data analysis algorithms improve with the adoption of 5G connectivity. All of these factors suggest that multimodal wearable recordings are here to stay, and could reveal new insights into long timescale dynamics of human health and disease, and seizure risk in epilepsy.

Medical grade wearable sensors have gained FDA approval for a variety of indications, including seizure detection.^38^ There have been a number of novel applications of commercial wearable devices,^39^ including identification of atrial fibrillation,^40^ and presymptomatic COVID-19 detection.^41^ Chronic ambulatory monitoring of human physiology gives new clinical relevance to chronobiology and chronotherapy, in which interventions are provided or behaviors adjusted to personalized time-varying models of disease. Epilepsy and drug resistant epilepsy are common,^42^ and antiseizure medications are often accompanied by side effects which limits quality of life.^43^ Chronotherapy may improve seizure control while minimizing therapy side effects and toxicity.^44^

Here, we tested the hypotheses that the homeostatic regulation of physiology fluctuates over multiscale cycles (circadian and multiday), that these long timescale dynamics can be measured by a multimodal research-grade wrist-worn device, and that the timing of electrographic seizures recur at a preferred phase of physiological cycles. All participants were monitored with an intracranial responsive neurostimulation (RNS) device that provides objective measures of IEA and electrographic seizure activity, and with a multimodal wearable research device from which we derived measures of heart rate (HR), beat-to-beat heart rate variability (HRV), accelerometry (ACC), temperature (TEMP), and tonic and phasic components of electrodermal activity (EDAt and EDAp) (Fig. 1). Time-frequency analyses were used to assess multiscale cycles in chronic brain and wearable recordings, and to evaluate for electrographic seizure phase locking to the recordings.

**Figure 1.**
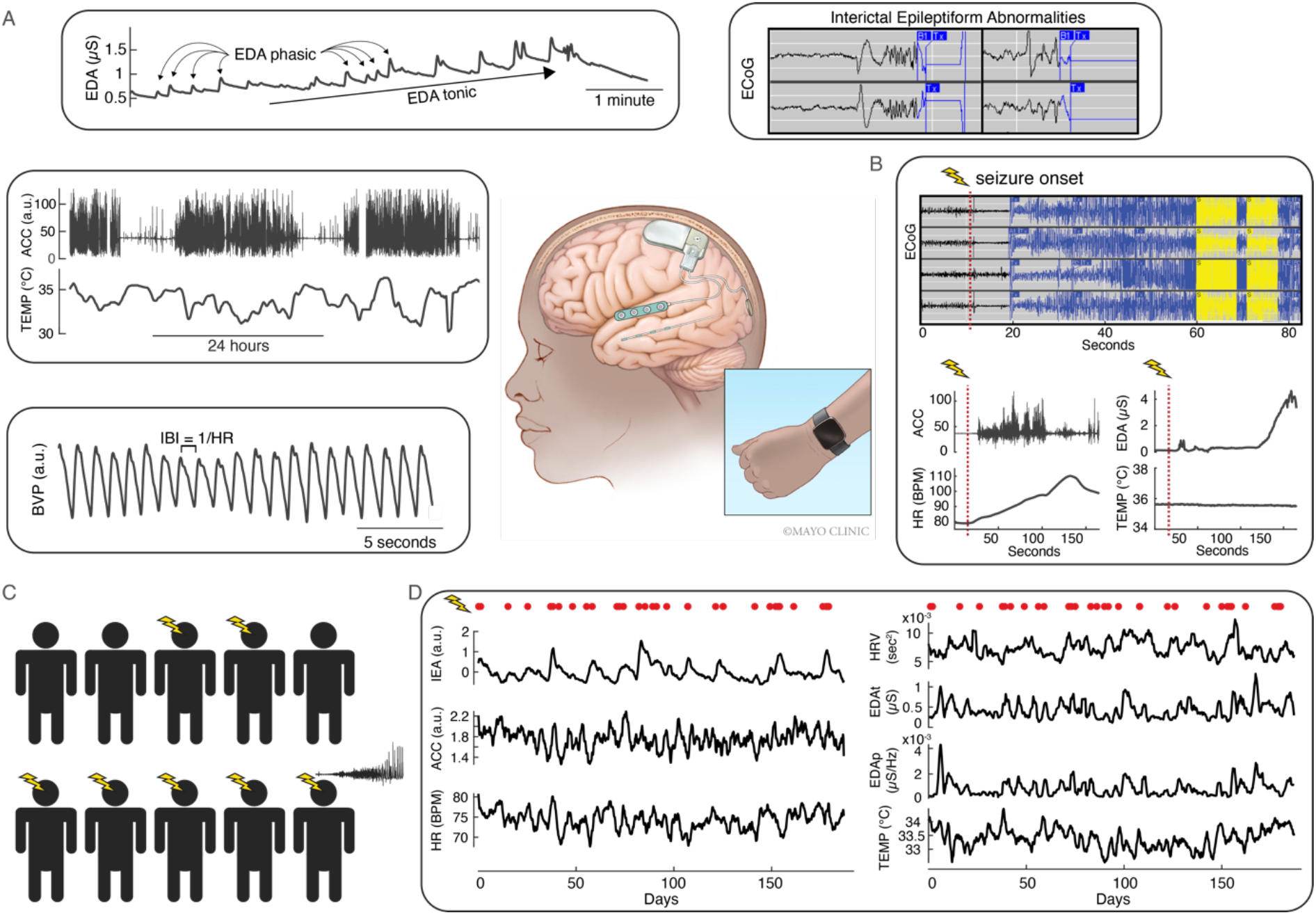
Chronic brain and wearable recordings. Unprocessed chronic brain and wearable device recordings. **A)** Brain and wearable recording device illustration, and representative interictal recordings. **B)** Concurrent brain and wearable ictal recordings (subject 3). **C)** Data from ten subjects were analyzed, seven of whom had reliable electrographic seizure detections (marked by lightning bolt). **D)** Concurrent chronic ambulatory brain and wearable recordings; 2-day moving average values; red dots mark seizure onset times. Interictal and chronic recordings are from subject 4. RNS = responsive neurostimulation. BVP = blood volume pulse. IBI = inter-beat-interval. HR = heart rate. EDA = electrodermal activity. ACC = accelerometry. TEMP = temperature. IEA = Interictal epileptiform activity.

## Materials and methods

### Subjects

Subjects with drug resistant epilepsy who were undergoing clinical treatment with an implanted response neurostimulation device (NeuroPace RNS®)^45^ were considered for enrollment in this observational cohort study of the feasibility of wearable devices to inform cycles of seizure risk in epilepsy. The RNS device had been placed and programmed per routine clinical care prior to enrollment in this study. Prescreening attempted to identify individuals with reliable electrographic seizure detections with the RNS brain monitoring system. Fourteen individuals provided signed informed consent and were enrolled in the study, and underwent concurrent monitoring with the RNS device and a multimodal wrist-worn sensor device (E4, Empatica Inc., Milan, Italy). Participants were provided with two wrist-worn devices which were swapped daily to have continuous recordings during device recharging and data download. Prior to this study, an epilepsy monitoring unit-based assessment of the E4 wearable device and external clock synchronization demonstrated clock drift of approximately 2 seconds per day. The RNS device has clock error on the order of 0.5-1 second. Both the brain implant and wearable device clocks are updated with each at-home patient data download, both which occurred daily to several times weekly, ensuring adequate temporal synchronization. Inclusion criteria for this work required at least 100 days of monitoring with the RNS and wearable devices. Subjects were recruited between November 2019 and February 2021 with monitoring and follow-up lasting up to one year. All subjects received their clinical care at Mayo Clinic. The study was approved by the Mayo Clinic Institutional Review Board.

### Interictal epileptiform activity and electrographic seizure detections

The RNS device provides constrained intracranial EEG (iEEG) recordings, and uses clinician defined detectors to quantify interictal epileptiform activity (IEA) and electrographic seizures (Fig. 1), as previously described.^4,46^ The hourly rate of IEA is recorded by the implanted device, and these data were used to assess cycles in IEA activity. The RNS device stores between 60 and 180 seconds of timeseries data (a ‘long-episode’ in NeuroPace terminology) when the seizure detector is triggered and can store up to approximately 10 minutes of timeseries data on the implanted device. ‘Long-episode’ recordings are overwritten in chronological order when on-device memory is full. This data is downloaded, approximately daily using an inductive telemetry device and patient tablet, with subsequent upload to cloud-based storage for clinician review. The device’s on-board memory is cleared after each upload. As previously described^46^ electrographic seizures were verified by a board certified epileptologist (N.M.G) and experienced reader (J.B.). Seizure analyses were limited to subjects for whom ‘long-episodes’ available for visual confirmation did not exceed the device’s storage capacity except rarely. Only visually reviewed ‘long-episode’ recordings were considered for seizure classification in this study (Fig. 1). At least 20 seizures were required per patient for analysis of seizure phase locking to brain and wearable recordings.

### Wearable sensor recordings

The wrist-worn Empatica E4 device provides measures of blood volume pulse (BVP) from which HR and HRV were calculated, 3-axis ACC, TEMP, and EDA from which tonic and phasic components were calculated. EDA, a measure of skin conductance, measures autonomic sympathetic arousal through autonomic innervation of sweat glands (Fig. 1).^47,48^

### Preprocessing wearable sensor recordings

Chronic ambulatory wrist-worn sensor recordings are susceptible to artifacts, and noisy and non-recording epochs were identified and removed from analyses using validated signal quality index measures.^49^ Briefly, HR and HRV signal quality was assessed by the variance in the peak-to-trough amplitude of the raw BVP signal assessed in 5 second non-overlapping blocks, after removing minor peaks (such as dicrotic notch). The peak-to-trough variance rejection threshold was determined by the knee-point (MATLAB function knee_pt.m, Dmitry Kaplan, 2012) in the sorted variance values (we found a cut-off of ½ the knee-point value provided the best balance of sensitivity and specificity for noisy epochs). Noisy epochs were removed. The ACC signal was calculated as the root-mean-squared value of 3-axis ACC recordings. ACC and TEMP data were removed for epochs in which the device was suspected to be off the body (TEMP outside of the range of 29-40 deg C). The EDA signal is prone to movement artifacts, and the signal quality index was defined as the rate of amplitude change in the raw EDA signal over 1 second epochs^49^ with the tenth percentile of extreme values removed, coupled with the BVP based signal quality assessment using ½ knee-point value noise threshold.

Heart rate was calculated from the raw BVP signal (*HR = 1/IBI*); (*IBI =* interbeat interval). Beat-to-beat HRV was defined as: *HRV =* (*IBI*_*i*+1_ − *IBI*_*i*_)^2^. The tonic and phasic components of the EDA signal were assessed independently. As previously described,^47,48^ the raw preprocessed EDA data were low pass filtered using a zero-phase shift 4^th^ order Butterworth filter with 0.045 Hz cutoff frequency to generate EDAt, while EDAp was equal to the signal of the 0.05 Hz to 0.25 Hz band of the continuous Morlet wavelet transform of raw preprocessed EDA data.

The denoised data were used to generate hourly average values for all wearable signals: ACC, HR, HRV, EDAt, EDAp, and TEMP. Hours in which at least thirty epochs (five seconds per epoch) were retained after denoising were used to calculate hourly average values, in non-overlapping one-hour blocks. The hourly rate of IEA was taken directly from the RNS system. Preprocessed TEMP values were used for subsequent analyses. Example 2-day moving average tracings from a single subject are shown in Fig. 1D, with 10 point smoothing.

### Data discontinuities

Data drops were filled using a 49-hour moving median interpolation (accounting for slow fluctuations in physiology), with additional circadian cycle correction using Pearson system random numbers drawn from the distribution of data corresponding to the same hour of the day (Pearson random numbers account for non-normal skew and kurtosis). Subjects had generally excellent adherence with device use (Table 1) and gaps in recording were uncommon. The entire recording duration was included for analyses of cycles in human physiology and seizure phase locking, to maintain consistency in record durations and seizure counts across the seven data channels.

**Table 1.** Subject characteristics. Intact recordings per channel lists the proportion of the entire recording duration for which there was data for hourly averaged brain and wearable physiology. R = right. L = left. Hc = hippocampus.

|  |  |  |  |  |  | Intact hourly averaged recordings |  |  |  |  |  |  |
| --- | --- | --- | --- | --- | --- | --- | --- | --- | --- | --- | --- | --- |
| Subject | Age | Gender | Lead location | Seizure count | Record (days) | IEA | ACC | HR | HRV | EDAt | EDAp | TEMP |
| 1 | 50s | F | R frontal strips | na | 144 | 0.92 | 0.90 | 0.89 | 0.88 | 0.89 | 0.89 | 0.90 |
| 2 | 40s | F | Bilateral Hc | na | 239 | 1.00 | 0.94 | 0.93 | 0.92 | 0.94 | 0.94 | 0.93 |
| 3 | 40s | F | L temporal neocortex | 24 | 200 | 1.00 | 0.93 | 0.93 | 0.93 | 0.93 | 0.93 | 0.92 |
| 4 | 30s | F | Bilateral Hc | 39 | 187 | 1.00 | 0.92 | 0.92 | 0.82 | 0.92 | 0.92 | 0.71 |
| 5 | 20s | F | Bilateral Hc | na | 208 | 1.00 | 0.97 | 0.97 | 0.91 | 0.97 | 0.97 | 0.97 |
| 6 | 40s | M | L frontal L insula | 20 | 270 | 1.00 | 0.90 | 0.89 | 0.79 | 0.89 | 0.89 | 0.89 |
| 7 | 50s | M | Bilateral Hc | 239 | 335 | 1.00 | 0.94 | 0.94 | 0.73 | 0.94 | 0.94 | 0.93 |
| 8 | 20s | F | Bilateral Hc | 150 | 269 | 0.90 | 0.69 | 0.69 | 0.61 | 0.69 | 0.69 | 0.69 |
| 9 | 20s | M | L temporal neocortex strips | 33 | 231 | 0.88 | 0.70 | 0.66 | 0.59 | 0.68 | 0.68 | 0.70 |
| 10 | 20s | F | L Hc L neocortex | 30 | 242 | 1.00 | 0.86 | 0.86 | 0.76 | 0.86 | 0.86 | 0.83 |
| Mean | 37 |  |  | 76 | 233 | 0.97 | 0.88 | 0.87 | 0.79 | 0.87 | 0.87 | 0.85 |

**Table 2.** Multiscale seizure risk categorization. Table corresponds to the results in Figure 7. Relative risk, high risk vs. low risk was defined as 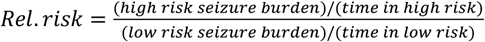. The last two rows list the total number of subjects included per signal, and the total number of phase-phase analyses per signal (more than 1 phase-phase assessment per signal per subject is possible); green, yellow, and red shading indicates high, moderate, and low subject/phase-phase analyses counts, respectively. IEA = interictal epileptiform activity. ACC = accelerometry. HR = heart rate. HRV = heart rate variability. EDAt = tonic electrodermal activity. EDAp = phasic electrodermal activity. TEMP = temperature.

|  | IEA | ACC | HR | HRV | EDA<sub>t</sub> | EDA<sub>p</sub> | TEMP | HR residual |
| --- | --- | --- | --- | --- | --- | --- | --- | --- |
| High risk: Time in | 0.08 | 0.08 | 0.08 |  | 0.04 | 0.07 | 0.09 | 0.07 |
| Seizure burden | 0.44 | 0.44 | 0.33 |  | 0.17 | 0.35 | 0.37 | 0.34 |
| Med. risk: Time in | 0.15 | 0.13 | 0.18 | 0.31 | 0.23 | 0.17 | 0.18 | 0.16 |
| Seizure burden | 0.29 | 0.26 | 0.38 | 0.54 | 0.42 | 0.35 | 0.39 | 0.31 |
| Low risk: Time in | 0.77 | 0.79 | 0.75 | 0.69 | 0.73 | 0.76 | 0.72 | 0.77 |
| Seizure burden | 0.27 | 0.30 | 0.29 | 0.46 | 0.42 | 0.30 | 0.24 | 0.35 |
| Rel. risk, H.R. v L.R. | 14.8 | 14.0 | 11.2 | nan | 7.7 | 11.7 | 12.6 | 10.5 |
| Subjects (n) | 5 | 2 | 3 | 2 | 3 | 3 | 4 | 5 |
| Phase-phase analyses (n) | 9 | 2 | 4 | 2 | 4 | 5 | 4 | 7 |

### Circadian and multiday cycles in epilepsy

#### Time-frequency analyses of brain and wearable recordings

Significant circadian and multiday cycles in chronic brain and wearable recordings were identified using a previously described method.^6^ Briefly, amplitude spectral density (ASD) plots were generated from the time-averaged continuous Morlet wavelet transform of hourly averaged brain and wearable recordings, for the identification of circadian and multiday cycles. A 1,000 trial simulation using normally distributed noise defined the 5^th^ percentile significance threshold. Significant cycles were defined as relative maxima in the ASD plots for circa-12-hour, 1-day, weekly (5-9 days), bi/tri-weekly (10-24 days), and monthly (25-35 days) cycles. Multiday groupings were defined in accordance with prior work describing group-level multiday chronotypes.^12^ At most one period was included per multiday category for analyses of seizure phase locking; the cycle of shortest period length was used when more than one maximum was present within a multiday category.

With cycles of interest identified for each brain and wearable channel, the filter-Hilbert analytic signal determined instantaneous measures of phase, frequency, and amplitude, as we have previously described,^6^ and similar to prior work.^4^ Filtering was performed with third-order zero-phase shift (non-causal) finite impulse response filters, with center periods, in units of days, of 0.3 to 2.0 incremented by 0.1, 2.5 to 10 incremented by 0.5, and 11 to maximum period length incremented by 1.0. Maximum period length was determined by the record duration and filter spread. The analytic signal was calculated with the Hilbert transform. Data within the cone of influence of boundary effects were excluded as previously described.^6^

#### Regression of behavioral covariates (ACC)

Behavioral activities can impact physiology measured by wearable sensors, and these behavior-induced changes may not be related to brain excitability or seizure propensity (i.e. heart rate change in response to exercise). We evaluated covariation between ACC and the other wearable signals to characterize the impact of behavior on wearable recordings. ACC was used as a proxy for behavior activities. The MATLAB Curve Fitting Tool was used to fit and regress the ACC signal from each wearable channel (see Supplementary information for regression model details). Goodness of fit was assessed by the coefficient of determination (R^2^). These analyses were performed on the 2-day moving average of all signals to prevent the circadian behavioral cycle from dominating the correlation. The residual wearable recordings after regression of correlated behavioral activity were evaluated for multiday cycles in epilepsy. Multiday cycle periods of interest were defined strictly as relative maxima in the ASD plot similar to prior work.^4^ Again, as described above, at most one cycle period was evaluated for circa-weekly, bi/tri-weekly, and monthly cycles.

### Seizure phase locking to cycles in epilepsy

The timing of each recorded electrographic seizure was assessed relative to the instantaneous phase of circadian and multiday cycles for each brain and wearable recording channel. This study relied on electrographic seizure detections from the RNS device. Prior work has demonstrated that endogenous cycles in human physiology may shift relative to clock time and vary around a central period tendency, especially in the absence of a Zeitgeber.^16,32,34,50^ To accommodate these cycle-to-cycle dynamics the Hilbert-transform analytic signal was calculated from the average bandpass composite centered at the period duration of interest (periods within center period +/- 25%), as previously described.^6^ Circular statistics (see Statistics section below) were used to characterize seizure phase locking to cycles in human physiology.

### Coherence of brain and wearable device recordings

Coherence provides a frequency band specific measure of the correlation between two signals. The magnitude squared coherence of brain and wearable recordings was evaluated at circadian and multiday cycle durations of interest (defined above) for each wearable channel. The association between brain-wearable coherence and seizure phase locking (resultant vector or R-value amplitude) was evaluated for multiday cycles using a linear regression model.

### Phase-Phase plots

Phase-phase plots were used to characterize the combined impact of circadian and multiday cycles on the timing of seizures, or in other words, the idealized seizure risk (non-causal zero-phase shift analyses). Phase-phase plots provide an intuitive representation of multiscale seizure risk by depicting together the phase data from different cycles for each seizure. Phase-phase plots were generated with surface fitting (surface linear interpolation) of 3-D histograms of seizure timing in phase-phase space. Seizure risk states (high, medium, low) were categorized by applying thresholds to phase-phase data (Fig. 6, Fig. 7). Risk thresholds were assigned such that approximately 5-10% of phase-phase space time was spent in the high risk state, approximately 10-20% in the medium risk state, and approximately 70-80% in the low risk state. Relevant features are the proportion of seizures that occur in each risk state and the proportion of time spent in each state. A relative risk metric was defined as:

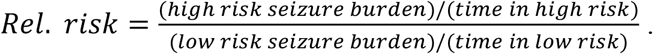

### Statistics

Circular statistics provide a framework for analyzing directional data, which is relevant for seizure phase locking to multiscale physiological oscillations. We refer readers to a recent review of cycles in epilepsy,^3^ which covers in detail the application of circular statistics in epilepsy. We have recently described the approach used here.^6^ All analyses were performed with MATLAB. Circular statistics were calculated with the MATLAB CircStat Toolbox.^51^ This package was used to generate circular histograms, and calculate the amplitude and phase angle of the resultant vector, or R-value, of seizure timing relative to the phase of physiological cycles. An R-value = 0 indicates that seizure timing has no phase preference for a given cycle; an R-value = 1 indicates that seizures recur at the same phase of an underlying physiological cycle. The Rayleigh test characterized the statistical significance of seizure phase locking to cycles or the degree of directional nonuniformity of seizure timing relative to the phase of cycles. Statistics were evaluated at the 0.05 significance level. All statistical tests were two-sided. Benjamini-Hochberg false discovery rate correction^52^ was applied to all circular statistics using n=7 recording channels. The association between brain-wearable coherence and seizure phase locking (R-value) was evaluated using a linear regression model with p-value determination.

### Data availability

Wearable data will be made available in 2023 on EpilepsyEcosystem.org. The analysis approach is described in detail above; MATLAB scripts are available from the author by reasonable request. All analyses were performed using MATLAB_R2020b, MathWorks® (mathworks.com).

## Results

### Subjects

Fourteen subjects were enrolled in the study, and 10 subjects met inclusion criteria, 7 of whom had reliable seizure detections. Subject characteristics are listed in Table 1. Four subjects had insufficient device adherence to meet inclusion criteria.

### Chronic brain and wearable recordings

Overall there was excellent participant adherence with the implanted and wearable devices (Table 1). In rare instances when a wearable device malfunctioned, this was identified through remote monitoring of regular data uploads and a new device was mailed to the participant.

### Multiscale cycles in human physiology

There was a high prevalence of ultradian (cycle frequency faster than daily), circadian, and multiday cycles across subjects and signals (Fig. 2). Prominent circadian cycles were seen in IEA, ACC, HR, HRV, EDAt, and TEMP, more so than EDAp. There was relatively greater amplitude of multiday cycles in IEA compared to wearable channels, however, significant multiday cycles were seen in all subjects for all wearable channels. Circa-weekly and biweekly cycles in particular were prominent in wearable recordings for some subjects.

**Figure 2.**
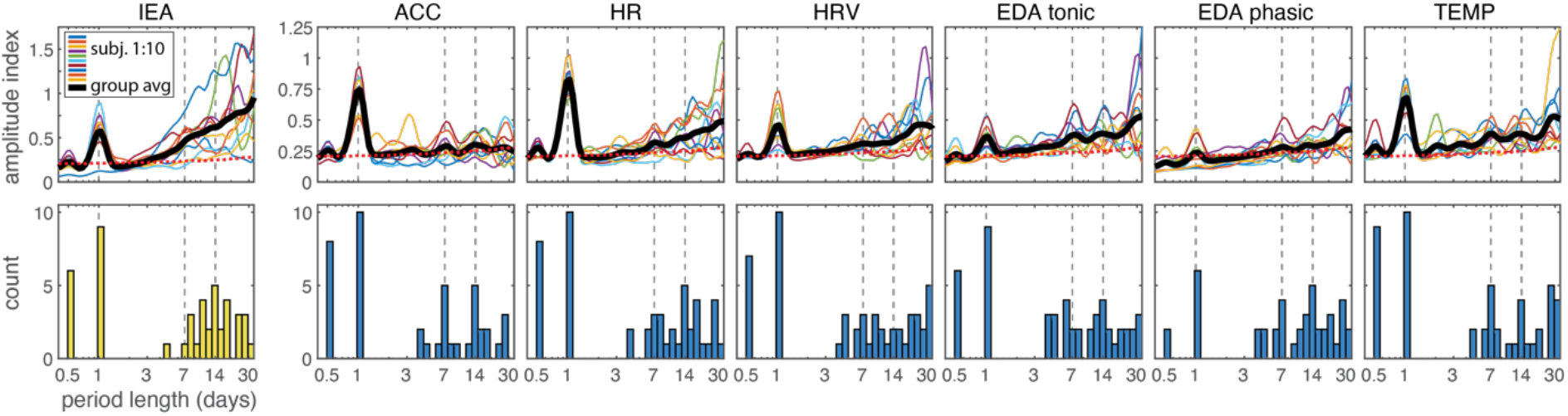
Amplitude spectral density. Amplitude spectral density of chronic brain and wearable device recordings. The top row presents the time averaged ASD for each channel across all subjects. The bottom panel shows the count of relative maxima for multiday cycles in the ASD above the 95^th^ percentile of normally distributed white noise (red dotted line). Vertical grey dashed lines mark daily, 7-day, and 14-day cycle periods. ASD = amplitude spectral density. IEA = interictal epileptiform activity. ACC = accelerometry. HR = heart rate. HRV = heart rate variability. EDAt = tonic electrodermal activity. EDAp = phasic electrodermal activity. TEMP = temperature.

### Circadian cycles and seizure risk in epilepsy

Circadian cycles were common across all recording channels and seen with ACC, HR, HRV, TEMP (n=10 subjects), IEA and EDAt (n=9 subjects), and EDAp (n=6 subjects). Daily cycles can be evaluated relative to fixed clock-time (Fig. 3A), or can be defined by approximately daily fluctuations in physiology as described in methods above (Fig. 3B). Figure 3B polar plots show the resultant vector of seizure phase locking to circadian cycles in physiology. By defining cycles by approximately daily changes in physiology, this method can accommodate drift in cycle length from day to day (i.e. variable weekday vs weekend schedules or shift work).

**Figure 3.**
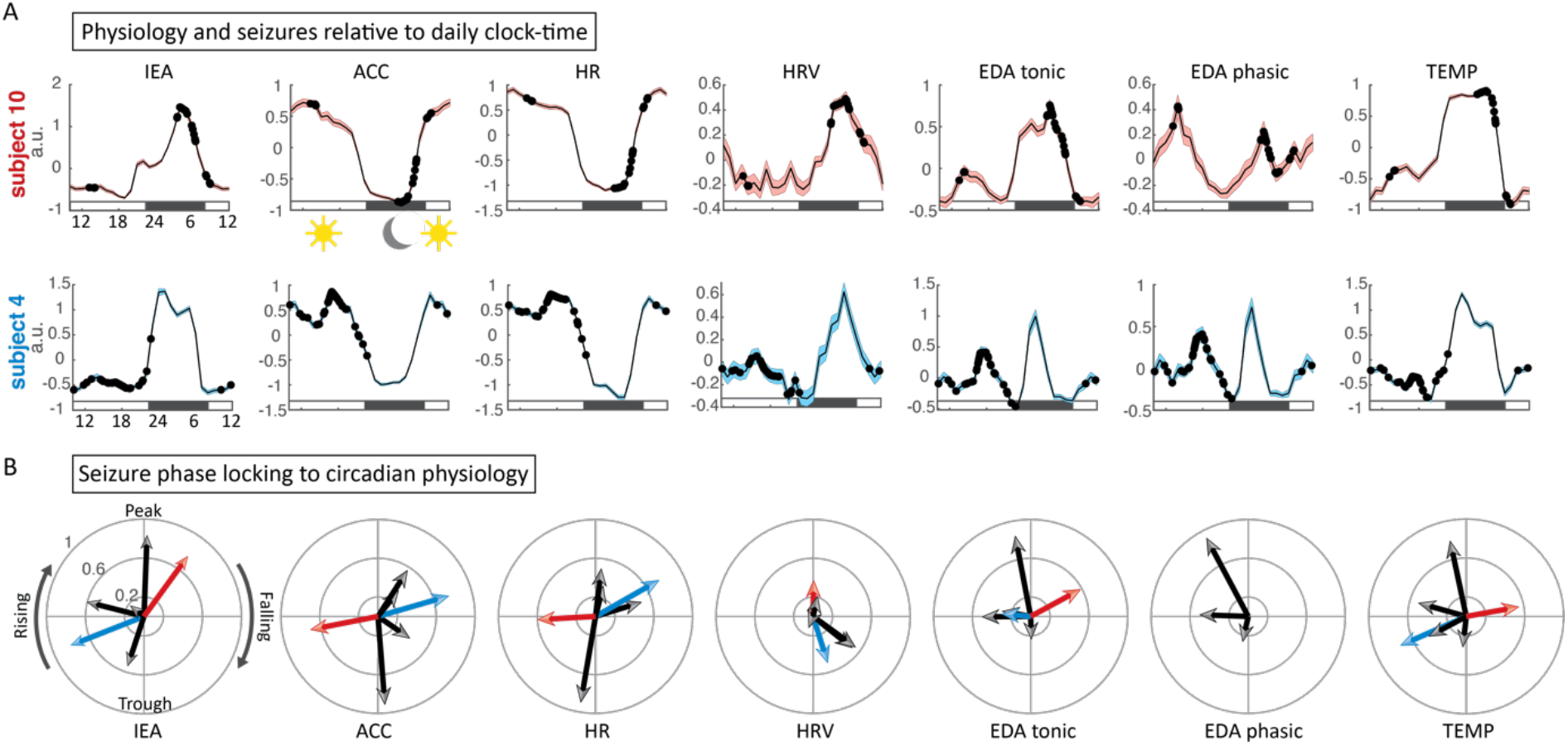
Circadian cycles of seizure risk. **A)** Daily average IEA and wearable recordings over the duration of monitoring, relative to clock-time. Error bars reflect standard error of the mean. Seizures relative to daily clock-time are marked by black circles. **B)** Resultant vector of seizure phase locking to circadian physiology. Only statistically significant resultant vectors are shown. Red and blue arrows correspond to the subjects in A) and black arrows correspond to the remaining subjects. IEA = interictal epileptiform activity. ACC = accelerometry. HR = heart rate. HRV = heart rate variability. EDAt = tonic electrodermal activity. EDAp = phasic electrodermal activity. TEMP = temperature.

Seizure phase locking to circadian cycles in brain and wearable recordings was common. Seven subjects had reliable electrographic seizure detections; three subjects had seizure phase locking to circadian EDAp cycles; six had seizure phase locking to circadian cycles for all other signals (Fig. 3B, Fig. 5B). There was no clear group level circadian phase preference for seizure timing, consistent with prior work using chronic brain recordings.

### Behavioral activity and wearable recordings

The correlation between wearable recordings and behavioral activity (ACC) is shown in Figure 4. The regression fit and coefficients of determination (R^2^ values) are shown in Fig 4A. HR had the strongest correlation with ACC, with median R^2^=0.32, while median R^2^ was <0.10 for each remaining wearable channel, (P=0.0025, two-sample *t*-test) (Fig. 4B). Cycles in epilepsy were assessed using the residual HR signal after regression of behavioral activity.

**Figure 4.**
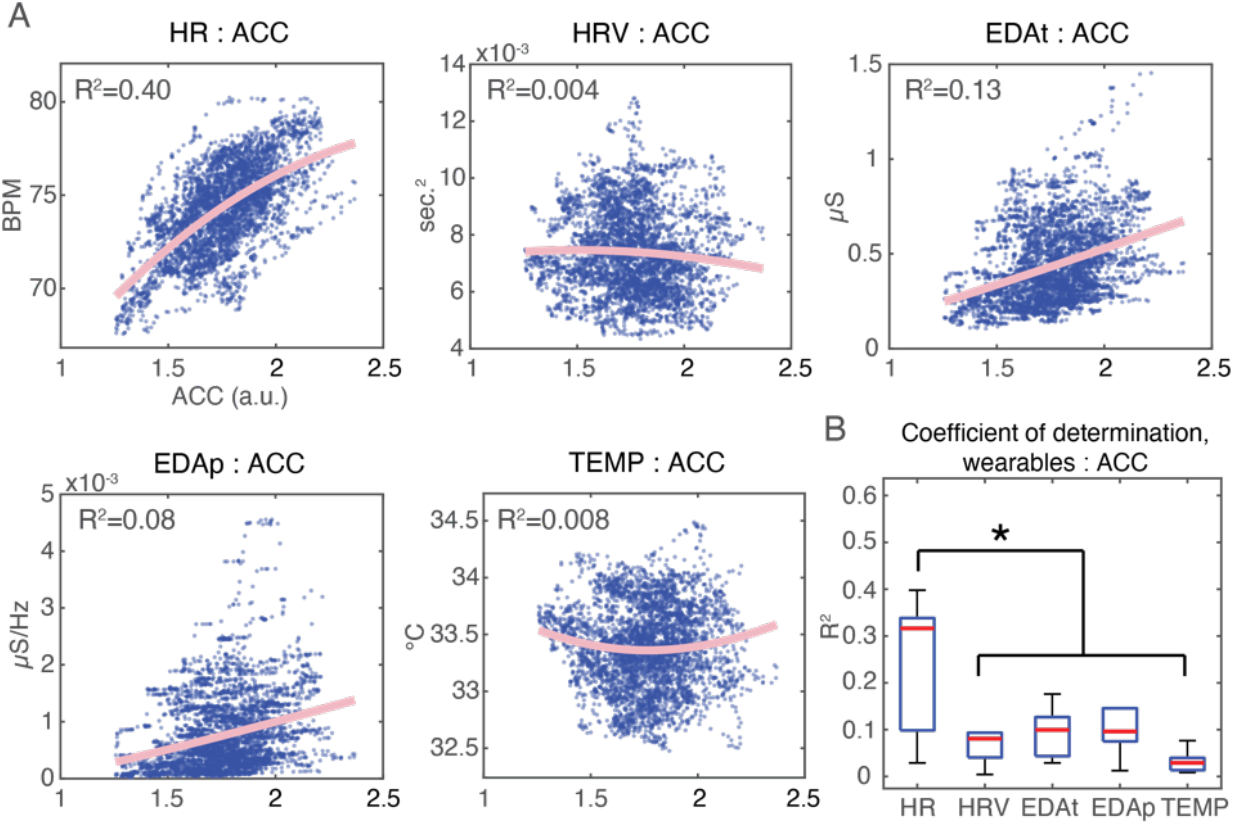
Wearable recordings and behavior. Correlation between wearable recordings and behavioral activity (ACC). **A)** Example scatter plots (subject 4) of wearable recordings relative to ACC, and linear model fit. **B)** Box plot of R^2^ values across all subjects for all wearable signals. * P=0.0025, two-sample *t*-test evaluating HR relative to the remaining channels; two-tailed. ACC = accelerometry. HR = heart rate. HRV = heart rate variability. EDAt = tonic electrodermal activity. EDAp = phasic electrodermal activity. TEMP = temperature. R^2^ = coefficient of determination.

### Multiday cycles and seizure risk

Multiday cycles were evaluated for all signals and subjects (Fig. 2). Seizure phase locking to multiday cycles was most prevalent in the IEA recordings (n=6 out of 7 subjects), followed by TEMP (n=5 subjects), EDAp and HR (n=4 subjects), and lastly ACC, HRV, and EDAt (n=3 subjects). Figure 5B shows the total number of multiday cycles with significant seizure phase locking across all subjects (evaluated for circa-weekly, bi/tri-weekly, and monthly cycles durations). The residual HR signal (with regression of behavioral factors (ACC)), when compared to the original HR, was notable for an increase from 4 to 6 subjects with seizure phase locking to multiday cycles, with an increase from 5 to 8 multiday cycles in total. Supplementary Table 1 provides subject specific data on clinical characteristics and seizure phase locking to multiday cycles counts.

**Figure 5.**
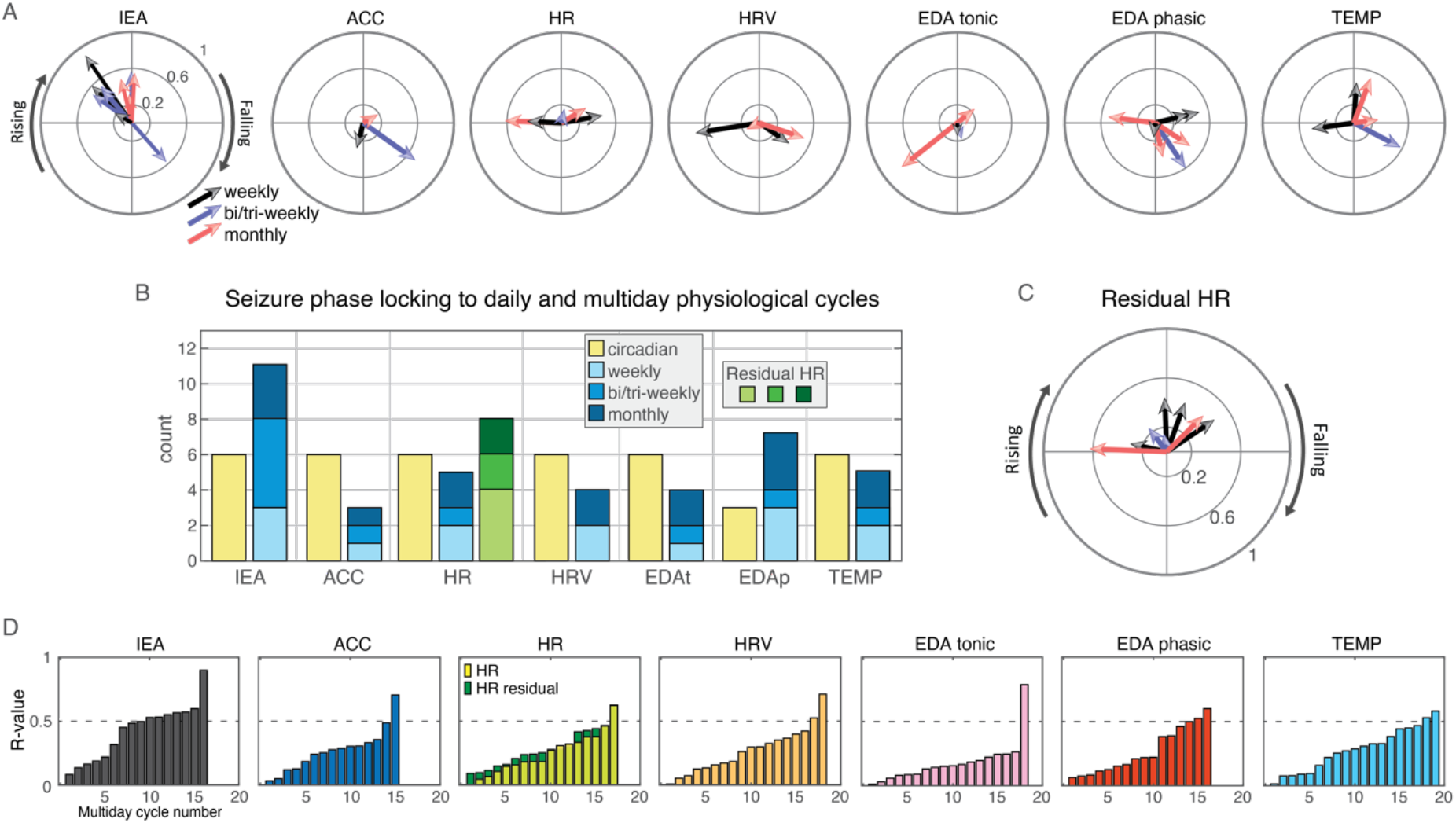
Multiday cycles of seizure risk. **A)** Polar plots of the resultant vector of seizure phase locking to multiday cycles of chronic brain and wearable recordings. **B)** Histogram showing the prevalence of significant seizure phase locking to circadian and multiday cycles. Seizure phase locking to residual HR after regression of behavioral activity (ACC) is marked in shades of green. **C)** Polar plot of the resultant vector of seizure phase locking to multiday cycles of residual HR. **D)** Sorted resultant vector amplitude (R-value) for seizure phase locking to multiday cycles. The HR plot shows seizure phase locking R-values for HR cycles and residual HR cycles. Plots show R-value for all significant peaks in amplitude spectral density plots, (not limited to cycles with significant seizure phase locking as in A)-C)). IEA = interictal epileptiform activity. ACC = accelerometry. HR = heart rate. HRV = heart rate variability. EDAt = tonic electrodermal activity. EDAp = phasic electrodermal activity. TEMP = temperature.

**Figure 6.**
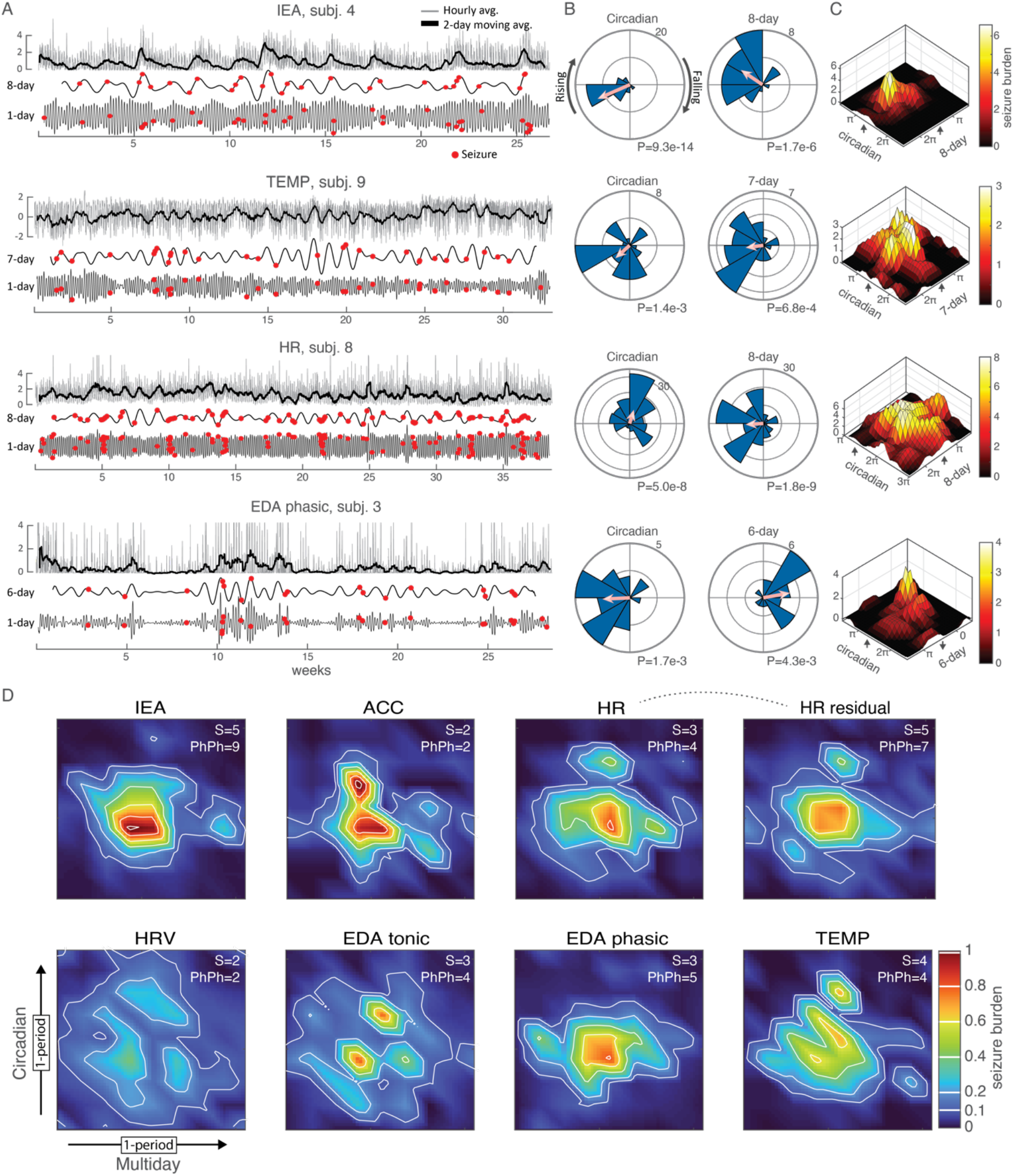
Multiscale cycles of seizure risk. **A)** Examples of chronic brain and wearable recordings, and below, circadian and multiday bandpass filtered tracings and seizure onset times. **B)** Corresponding polar histogram plots, with the R-value shown as a pink arrow. The number on the outer ring is seizure count for the histogram; for R-value amplitude, the outer ring = 1. **C)** Corresponding phase-phase plots show seizure counts with respect to circadian and multiday cycles. ∏ = cycle trough, 0 and 2∏ = peak, and ↑ and ↓ = rising or falling phase. **D)** Group averaged phase-phase plots for all significant circadian and multiday cycles. Phase-phase plots were normalized to the total number of seizures per subject prior to averaging so high seizure count subjects did not drive group results. Given subject specific phase preferences the median circadian and multiday phases were centered prior to averaging. The scale of the colormap has a fixed proportional scale relative to the total seizure count for each channel, for direct comparisons between recording channels. The top right inset script is the number of subjects (S) and the number of phase-phase analyses (PhPh) included in the group plot (cycles with significant seizure phase locking). IEA = interictal epileptiform activity. ACC = accelerometry. HR = heart rate. HRV = heart rate variability. EDAt = tonic electrodermal activity. EDAp = phasic electrodermal activity. TEMP = temperature. e = power of 10 (1e2 = 1×10^2^).

**Figure 7.**
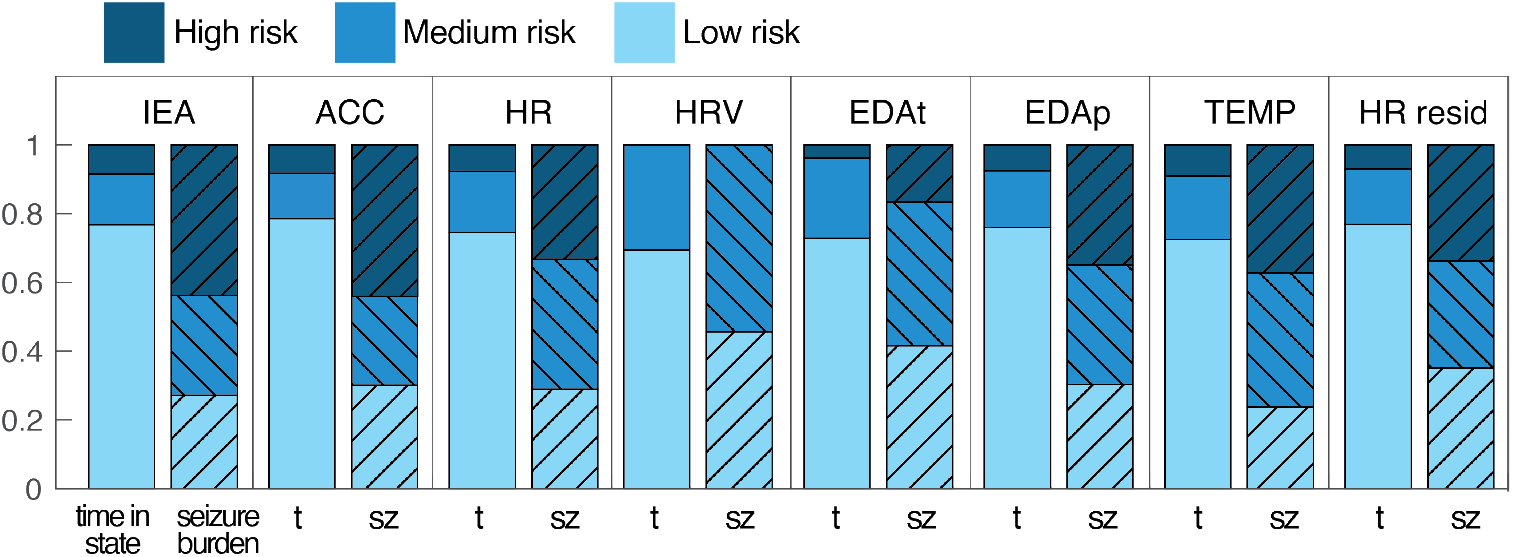
Multiscale seizure risk categorization. These results are derived from the phase-phase analyses shown in Figure 6. ‘Time in state’ is the proportion of the total phase-phase space categorized into that state. The time in high risk + medium risk + low risk = 1. ‘Seizure burden’ of a risk state is the proportion of all seizures that occur within that risk category. ‘Seizure burden’ high risk + medium risk + low risk = 1. IEA = interictal epileptiform activity. ACC = accelerometry. HR = heart rate. HRV = heart rate variability. EDAt = tonic electrodermal activity. EDAp = phasic electrodermal activity. TEMP = temperature.

There was a group level preference for seizure phase locking to the rising phase of multiday IEA cycles (with one outlier), as has been shown previously (Fig. 5A)^3,4^. Group level preferences were less evident in wearable recordings. There was a group level preference for seizure phase locking to the peak (late rising/peak/early falling) phase of the residual HR signal (Fig. 5C).

Figure 5D shows the R-value amplitude of seizure phase locking to all multiday cycles across recording channels (again, limited to one cycle per circa-weekly, bi/tri-weekly, and monthly categories). R-value amplitudes were generally larger for IEA than wearable signals. Seizure phase locking to the residual HR signal has increased R-value amplitudes compared to raw HR data, with one additional multiday cycle in total.

### Coherence between wearable and brain recordings

The magnitude of seizure phase locking (R-value) was evaluated relative to the degree of coherence between cycles in wearable recordings and IEA (Supplementary Fig. 2). There were high levels of coherence between wearable and brain recordings for circadian cycles (median coherence of IEA and wearable signals: ACC=0.89, HR=0.89, HRV=0.81, EDAt=0.75, EDAp=0.60, TEMP=0.83). For multiday cycles, a regression model of wearable to IEA coherence and seizure phase locking R-value did not demonstrate a consistent association between coherence and R-value; the linear regression model fit P-value > 0.05 for all signals. The linear fit model evaluating coherence and seizure phase locking to multiday cycles had a slope of 0.24, R^2^=0.10, P=0.21 for residual HR, compared to slope of 0.075, R^2^=0.0085, P=0.73 for the original HR signal.

### Multiscale cycles and seizure risk

The association between multiscale (circadian and multiday) cycles of human physiology and seizure risk is displayed in Figure 6. Figure 6A shows circadian and multiday cycles in chronic brain and wearable recordings from four subjects. Circadian cycles are evident in the high frequency component of the hourly average tracing, while multiday fluctuations in physiology are apparent in the 2-day moving average. Circadian and multiday bandpass filtered data is shown with overlayed seizure timestamps. Polar plots show the degree of seizure phase locking to circadian and multiday physiological cycles (Fig. 6B). Seizure risk is greatest during periods of cooccurrence of the high risk phases of circadian and multiday cycles (Fig. 6C). Figure 6D shows the group averaged phase-phase plots for all significant circadian and multiday cycles. The number of subjects, and number of phase-phase analyses included for each channel are listed in the top corner of each panel.

Phase-phase analyses provide a representation of the temporal distribution of seizures relative to the phase of multiscale cycles. Figure 6D shows a continuum of seizure risk levels indicated by the seizure burden color scale. Alternatively, phase-phase space data can be categorized into discrete seizure risk states, such as low risk, medium risk, and high risk. To illustrate the point, high risk (>0.4), medium risk (≥0.15 and ≤0.4) and a low risk (<0.15) thresholds were naively assigned for phase-phase analyses, and measures of the relative seizure burden and relative time in a risk state were calculated (Fig. 7). Risk thresholds were constant across all signals. For true seizure forecasting applications risk category thresholds could be optimized on an individual basis.

An ideal model of seizure risk would 1) capture a large proportion of seizures in the high risk state, which would occupy a small proportion of time, 2) have a low number of seizures occur in the low risk state (false negatives), and the low risk state would consume a large proportion of total time, and 3) be widely applicable for most individuals with epilepsy (for us, the proportion of subjects who were candidates for phase-phase assessment). IEA provided the best performance with respect to high/low risk discrimination and subject inclusion, followed by residual HR, followed by TEMP, HR and EDAp, followed by ACC with good performance but few subjects, and finally EDAt and HRV with poor performance and few subjects.

## Discussion

This work demonstrates that circadian and multiday cyclical changes in noninvasive measures of physiology are common in people with drug resistant focal epilepsy, and that seizures occur at preferred phases of these cycles for many people. Circadian and multiday cycles of seizure risk were seen across wearable recording channels, which included ACC, HR, HRV, EDAt, EDAp, and TEMP; multiscale cycles were present in concurrent chronic brain recordings of IEA, consistent with prior work.^3,4^ This work used a unique dataset of ultra-long-term ambulatory recordings from a wrist-worn device and a clinical RNS brain implant, containing over 2,300 days of recordings and 535 electrographic seizures from ten participants, seven of whom had reliable electrographic seizure detections. Every subject had seizure phase locking to multiday cycles for at least one wearable channel. Seizure phase locking to multiday cycles was most common for the HR and EDAp (n=4 out of 7 subjects each), and TEMP (n=5). Seizure phase locking to residual HR (HR after regression of behavioral covariates) multiday cycles had better performance and seizure phase locking was seen in six subjects. In comparison, six subjects had seizure phase locking to multiday IEA cycles. To the best of our knowledge this is the first study to assess long timescale cyclical changes in seizure risk with a multimodal wearable device, with concurrent brain recordings and objective electrographic seizure detections. These findings suggest that multimodal wearable sensors can inform long timescale cycles of seizure risk and highlights the importance of chronobiology in epilepsy.

Seizure phase locking to circadian cycles was common across brain and wearable recording channels (less so for EDAp compared to other signals) (Fig. 3). The suprachiasmatic nucleus^53^ of the hypothalamus—the master pacemaker—organizes and synchronizes a complex set of circadian physiological changes that operate at the level of gene and protein expression,^54^ and culminate in daily changes in endocrine,^55^ immune,^56^ metabolic,^31^ autonomic,^57^ and brain^17,58^ function, and behavioral^59^ (sleep/wake) state. Daily Zeitgebers from the earth’s rotation (natural light exposure; ambient temperature) maintains entrainment of an individual’s circadian cycle with the celestial day and prevents free-running drift in the cycle period.^50^ Circadian changes in epilepsy, including changes in interictal epileptiform discharges and seizure risk, are well known,^30^ and circadian chronotypes have been described in prior work on a large cohort of individuals monitored with the RNS device.^12^ While the relative importance of the circadian cycle^60^ vs. sleep/wake state^61^ needs further study, the ability of wearable devices to track circadian changes in physiology can allow for seizure risk tracking even in the absence of causal links between wearable recordings and brain excitability. The high degree of circadian coherence between wearable and brain recordings (in particular ACC and HR; Supplementary Fig. 2), further suggests wearables can effectively track circadian cycles of seizure risk, and accommodate to changing sleep/wake patterns (i.e. shift workers; changing sleep habits between weekends and weekdays) in contrast to fixed clock-time based cycles.

This work is particularly notable for the presence of seizure phase locking to multiday cycles in wearable recordings. This finding in wearable recordings is similar to seizure phase locking to multiday cycles in brain recordings as noted in this study and in prior work.^3,4,6,7,13^ It is perhaps unsurprising that seizure phase locking to multiday cycles was mostly reliably seen for IEA (Figs. 5), however, the benefits of accessibility and low cost of noninvasive wearable monitoring in comparison to invasive brain recordings is self-evident. Of the wearable channels, seizure phase locking to multiday cycles was most common for the residual HR channel, as well as for the EDAp, original HR, and TEMP channels (Fig. 5).

The physiological signals measured by the wearable device in this study can be impacted by volitional behavioral activities, which may be uncorrelated with seizure risk. A linear model was used to assess the degree of correlation between ACC (a proxy for behavior activities) with each wearable channel. HR was significantly more correlated with ACC than the remaining wearable channels (Fig. 4). A residual HR channel (HR after linear regression of the ACC signal) was included in analyses of multiday cycles. Notably, seizure phase locking to multiday cycles increased with residual HR (n=6 out of 7 subjects) compared to the original HR channel (n=4 of 7 subjects) (Fig. 5B, C, D). Additionally, there was a group level preference for seizure phase locking to the peak (late rising/peak/early falling) phase of the residual HR cycle (Fig. 5C). This might indicate that periods of increased autonomic arousal (reflected in peak residual HR activity) are associated with times of heightened brain excitability and seizure risk. The proportion of subjects with seizure phase locking to multiday cycles of residual HR is somewhat greater than prior work^21^ and may relate to differences in technique (original HR results were similar to prior work).

Each seizure occurrence can be described relative to the instantaneous phase of both circadian and multiday cycles to characterize the interplay of multiscale cycles on seizure risk (Fig. 6D, Fig. 7). Performance was assessed by 1) the relative risk of seizures in high, medium, and low risk states, 2) the proportion of seizures that occur in each risk state (ideally, a high proportion of seizures occur in the high risk state, while few seizures occur in the low risk state), 3) the proportion of time in each risk state, and 4) the number of subjects for whom cycles based analyses were applicable (Fig. 6D, Fig. 7). The best performance was achieved by IEA, followed by residual HR, followed by TEMP, EDAp, and HR, followed by ACC with poor subject counts but reasonable classification performance, and finally EDAt and HRV with poor subject counts and poor classification performance.

Interactions between brain excitability, autonomic arousal, various mechanisms of body homeostasis (endocrine, metabolic, immunologic), and behavior may influence long timescale fluctuations in wearable recordings and seizure timing. Further investigations of potential mechanisms are needed. An assessment of the association between IEA and wearable coherence and seizure phase locking to multiday cycles was not significant (Supplementary Fig. 2). The complexity of wearable recording, subject to various homeostatic and behavioral factors, is a challenge for direct comparisons of brain and wearable recordings. Multimodal sensors and models that control for behaviorally driven changes in physiology may improve seizure risk characterization, as seen with the residual HR channel.

Activation of several brain regions has been implicated in the regulation of autonomic arousal, including the amygdala,^23^ and the anterior cingulate, insula, thalamus, and regions of the prefrontal cortex,^62^ supporting the idea that changes in brain excitability or network activity may be associated with autonomic arousal. Cardiac function and EDA are under direct autonomic regulation. EDA is a measure of sympathetic arousal,^47,48^ and changes in sympathetic arousal, a measure of cognitive and physical stress, may impact seizure risk. Furthermore, changes in sleep patterns and sleep deprivation may impact sympathetic arousal^63^ and cortical excitability,^64^ and sleep deprivation is a known seizure risk factor.^65^ Some work suggests divergent mechanisms for the regulation of EDAp vs. EDAt,^62^ and artifactual drift in the EDAt signal is a concern for long recording periods.^47^ These factors may underpin differences in the EDAt and EDAp channels, with EDAp being the more meaningful feature for multiday seizure risk in this work.

Seizure phase locking to multiscale cycles in HR corroborates recent findings from a long-term study of seizure risk and heart rate cycles using patient reported seizure diaries and a fitness watch.^21^ Our work strengthens the finding with electrographic confirmation of seizures from chronic brain recordings, and the use of a multimodal wearable device. Prior work has established that ictal and interictal heart rate changes are common in epilepsy,^22,66^ however, only recently have the tools been available to assess for seizure cycles over ultra-long timescales. The increased efficacy of closed-loop vagus nerve stimulation (VNS) based on heart rate change further supports an important brain-heart or brain-autonomic arousal mechanism in epilepsy.^67^ More work is needed to characterize the interplay of behavior, brain excitability, and autonomic arousal to better extract a seizure risk signal from chronic wearable recordings.

The good performance of the TEMP signal is intriguing. Prior work has shown that surface TEMP and core TEMP are both involved in the subjective sensation of comfort and drive behavioral responses, while core TEMP is the primary driver of autonomic and metabolic homeostatic changes.^68^ The degree of similarity between surface and core TEMP is thus an important factor when considering the association of surface TEMP with autonomic and metabolic activity. Surface TEMP may be less prone to behavioral influences than HR. These results motivate future investigations of multiscale fluctuations in cutaneous surface TEMP and seizure risk.

Chronic ambulatory monitoring poses challenges that are relevant to this study. Chronic ambulatory wearable recordings are prone to artifacts, and EDA in particular is susceptible to movement artifacts and long-term drift in the signal.^47^ This work relied on previously validated^49^ signal quality indices to identify and remove noisy epochs. The long timescale changes of interest in this work allowed for relatively stringent signal quality controls, and the large datasets and hourly averaged signals further limit the impact of random artifacts. The use of objective electrographic seizure detections from a clinical responsive neurostimulation device,^45^ rather than patient reported seizure diaries avoids the unreliability of patient reported diaries,^69^ but does not distinguish electrographic vs. electroclinical seizures.

This is a relatively small cohort, however, the long recording durations bolster the reliability of results. Larger cohorts are needed to characterize the generalizability of these findings and to investigate the impact of seizure onset location on cycles in epilepsy (our cohort was enriched for temporal lobe epilepsy). False discovery rate correction was performed to account for multiple statistical comparisons. This retrospective analysis of long timescale seizure dynamics, consistent with prior efforts in the field,^3,4,6,12-14,21^ used zero-phase shift noncausal analyses, and efforts are underway for the application of cycles based seizure forecasting to prospective trials. The assessment of seizure risk in epilepsy may be improved further by combining long timescale cycles based seizure risk characterization with an acute seizure forecast layer, which has shown early promise with multimodal wearable devices.^46^ More work is needed to optimize the performance of multimodal wearable sensors, however, the results from our straightforward approach are encouraging. Even if multimodal wrist-worn devices are unable to attain the performance of a brain implant, the ease of implementation, lower cost, broad applicability, and patient preference for a wrist-worn form factor^49^ make this a critical area of study. Finally, future efforts should assess the utility of combining wearable and minimally invasive sub-cutaneous EEG^70^ or intracranial EEG recordings.^69^

This work provides evidence that long timescale cycles in noninvasive measures of human physiology are common in epilepsy, and that for many individuals, seizures occur at preferred phases of these cycles. Improvements in medical and consumer wearable devices will likely lead to wider adoption over time and advance the study of the chronobiology of epilepsy, and of health and disease generally.

## Data Availability

Wearable data will be made available in 2023 on EpilepsyEcosystem.org. The analysis approach is described in detail above; MATLAB scripts are available from the author by reasonable request.

## Abbreviations

ACC: accelerometry
ASD: amplitude spectral density
BVP: blood volume pulse.
EDAp: phasic electrodermal activity
EDAt: tonic electrodermal activity
HR: heart rate
HRV: heart rate variability
IBI: interbeat interval
IEA: interictal epileptiform activity
iEEG: intracranial EEG
RNS: responsive neurostimulation
TEMP: temperature

## Acknowledgements

This research was supported by the Epilepsy Foundation Epilepsy Innovation Institute My Seizure Gauge; NIH grants UH3-NS95495 & R01-NS09288203; American Epilepsy Society Research & Training Fellowship for Clinicians (N.M.G.); National Science Foundation grant CBET-2138378 (M.N.). The authors acknowledge technical and administrative support from Sherry Klingerman CCRP.

## Author contributions

Conceived and designed the work: N.M.G., B.H.B, D.R.F., M.P.R., G.A.W.

Acquired, analyzed, or interpreted data: N.M.G., T.P.A, M.N., B.J., G.A.W., B.H.B. All authors drafted or substantively revised the manuscript.

## Additional information

Supplementary information is available online.

**Correspondence** and requests for materials should be addressed to N.M.G.

