## Supplementary Info for "Multimodal wearable sensors inform cycles of seizure risk"

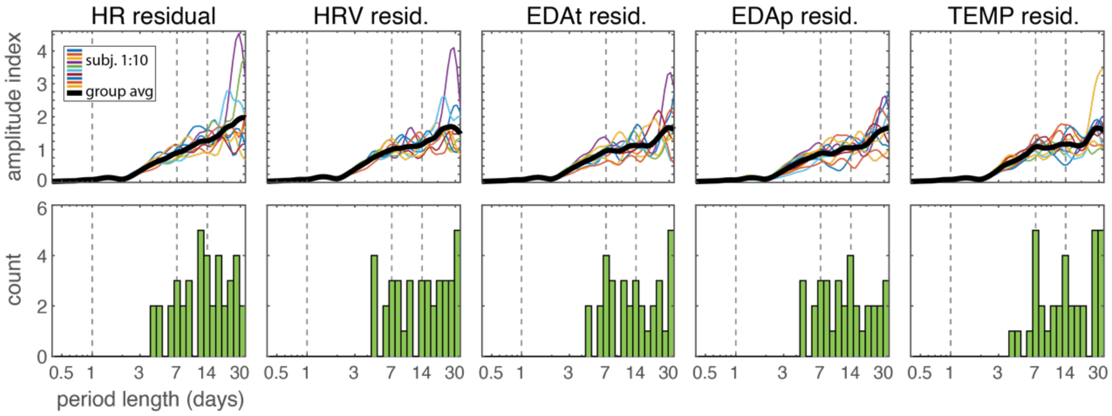


**Supplementary Figure 1 Amplitude Spectral Density.** Amplitude spectral density of residual wearable channels after regression of behavioral activity (ACC channel). The bottom panel shows the count of relative maxima for multiday cycles (subjects were limited to at most one cycle for each category of circa-weekly, bi/tri-weekly, and monthly cycles). Vertical grey dashed lines mark daily, 7-day, and 14-day cycle periods. HR = heart rate. HRV = heart rate variability. EDAt = tonic electrodermal activity. EDAp = phasic electrodermal activity. TEMP = temperature.

**Supplementary Table 1 Seizure phase locking to multiday cycles**

| | | | | | | Seizure phase locking to multiday cycles | | | | | | | |
|---|---|---|---|---|---|---|---|---|---|---|---|---|---|
| **Subject** | **Age** | **Gender** | **Lead location** | **Seizure count** | **Record (days)** | **IEA** | **ACC** | **HR** | **HRV** | **EDAt** | **EDAp** | **TEMP** | **HR residual** |
| 1 | 50s | F | R frontal strips | na | 144 | | | | | | | | |
| 2 | 40s | F | Bilateral Hc | na | 239 | | | | | | | | |
| 3 | 50s | F | L temp neocortex | 24 | 200 | 1 | 0 | 0 | 0 | 0 | 1 | 0 | 1 |
| 4 | 30s | F | Bilateral Hc | 39 | 187 | 2 | 0 | 0 | 0 | 0 | 0 | 0 | 1 |
| 5 | 20s | F | Bilateral Hc | na | 208 | | | | | | | | |
| 6 | 40s | M | L frontal L insula | 20 | 270 | 1 | 0 | 1 | 2 | 0 | 2 | 1 | 1 |
| 7 | 50s | M | Bilateral Hc | 239 | 335 | 2 | 1 | 1 | 1 | 1 | 2 | 1 | 1 |
| 8 | 20s | F | Bilateral Hc | 150 | 269 | 2 | 1 | 2 | 1 | 2 | 0 | 1 | 3 |
| 9 | 20s | M | L temp neocortex strips | 33 | 231 | 0 | 0 | 0 | 0 | 0 | 0 | 1 | 0 |
| 10 | 20s | F | L Hc L neocortex | 30 | 242 | 3 | 1 | 1 | 0 | 1 | 2 | 1 | 1 |
| Mean | 37 | na | na | 76 | 233 | 1.57 | 0.43 | 0.71 | 0.57 | 0.57 | 1.00 | 0.71 | 1.14 |
| Count | | | | | | 6 | 3 | 4 | 3 | 3 | 4 | 5 | 6 |

Subject characteristics and seizure phase locking to multiday cycles. Subjects were limited to at most one cycle for each category of circa-weekly, bi/tri-weekly, and monthly cycles.

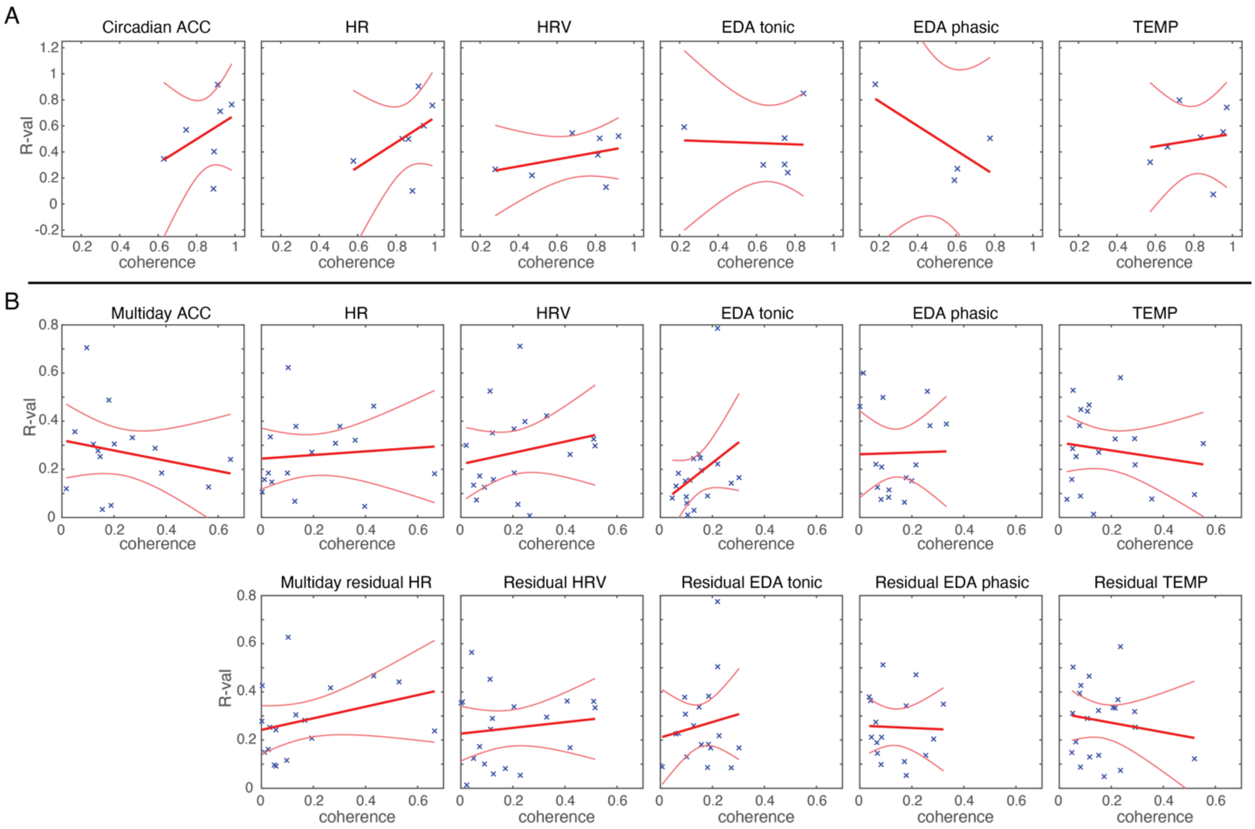


**Supplementary Figure 2 Seizure phase locking and IEA-wearable coherence**. Seizure phase locking resultant vector (R-value) amplitude relative to the coherence between wearable and IEA cycles, assessed for A) circadian and B) multiday cycles. The bold red line marks the linear regression fit with superimposed 95$^{th}$ percentile confidence bounds. There was not a significant association between coherence and seizure phase locking value for circadian or multiday cycles for any channels. The linear fit model evaluating seizure phase locking to multiday cycles had a slope of 0.24, $R^2$=0.10, P=0.21 for residual HR, compared to slope of 0.075, $R^2$=0.0085, P=0.73 for the original HR channel. IEA = interictal epileptiform activity. ACC = accelerometry. HR = heart rate. HRV = heart rate variability. EDAt = tonic electrodermal activity. EDAp = phasic electrodermal activity. TEMP = temperature. $R^2$ = coefficient of determination.

### Behavioral activity and wearable recordings

The MATLAB Curve Fitting Tool was used to fit and regress the ACC signal from each wearable channel. The Curve Fitting Tool regression model was:

$$f(ACC) = a * \sin(ACC - \pi) + b * (ACC - 10)^2 + c$$
